# Plasma proteomic signatures of preclinical Alzheimer’s disease biomarkers and memory in clinically unimpaired older adults

**DOI:** 10.1101/2025.10.22.25338569

**Authors:** Alexandra N Trelle, Karly A Cody, Tran T Nguyen, Joseph R Winer, Skylar Weiss, Isha Sai, Divya Channappa, Justin Mendiola, Amal Al-Rajhi, Keerthana Raghuraman, Sharon J Sha, Edward N Wilson, Tony Wyss-Coray, Anthony D Wagner, Holden T Maecker, Elizabeth C Mormino

## Abstract

**Background:** Multianalyte plasma proteomic panels that can accurately detect initial AD pathology in preclinical populations and simultaneously measure related biological processes relevant for disease risk are critical for advancing early detection and prognosis.

**Methods:** Using the NULISAseq CNS panel, we measured plasma from 193 clinically unimpaired (CU) older adults enrolled in the Stanford Aging and Memory Study (SAMS). We evaluated correspondence of core AD-relevant biomarkers Aβ42, Aβ40, pTau217, pTau181, GFAP, NfL, Aβ42/Aβ40, and pTau217/Aβ42 measured using NULISAseq and established Lumipulse immunoassays. ROC curve analyses compared the accuracy of these biomarkers for detecting Lumipulse CSF Aβ-positivity across platforms. Linear models were applied across 124 NULISAseq proteins to examine associations with common AD risk factors, including age, female sex, and *APOE-*ε4, as well as with biomarkers CSF Aβ42/Aβ40 and pTau181, 18F-PI2620 Tau PET, and memory. Fold change differences in NULISAseq proteins as a function of CSF Aβ (A+) and pTau181 (T+) status were examined using Wilcoxon rank-sum tests.

**Results:** Moderate to high correlations were observed between NULISAseq and Lumipulse AD plasma biomarkers. Across platforms, plasma pTau217/Aβ42 exhibited the highest performance in discriminating CSF A+ (NULISAseq AUC: 0.940, 95%CI: 0.885-0.995; Lumipulse AUC: 0.907, 95%CI: 0.849-0.966). Age and sex were associated with differential expression of NULISAseq targets linked to neurodegeneration, microglial activation, and inflammation. CSF A+ was associated with fold change differences in Aβ42, pTau217, pTau231, pTau181, and GFAP, while CSF T+ was additionally associated with increases in TREM1, TIMP3, SAA1, and S100A12. When stratified by AT groups, A+T-exhibited lower Aβ42 and elevated pTau217 compared to A-T-, whereas A+T+ exhibited elevated pTau231 and pTau181 compared to A+T-. Temporal cortex tau was positively associated with NULISAseq pTau217, pTau231, pTau181, and pTau217/Aβ42. Memory function was negatively associated with pTau isoforms and PRDX6, YWHAZ, ENO2, ARSA, CHI3L1, CXCL8, and FCN2. These associations remained when controlling for pTau217 and restricting to A-CU, suggesting these targets may represent AD-independent biological pathways relevant for memory function.

**Conclusions:** NULISAseq immunoassay-based multiplexing accurately detects AD pathology among CU older adults and identifies multiple biological pathways related to early biomarker abnormality and memory function that may become dysregulated in preclinical AD.

## Introduction

Effective diagnosis, treatment, and prevention of Alzheimer’s disease (AD), particularly in the era of disease-modifying therapeutics, relies critically on the availability of accessible and scalable biomarkers that can accurately detect AD pathology in vivo. The recent development of blood-based biomarkers that can accurately detect cerebral amyloid-β (Aβ) pathology is transforming the landscape of AD research and clinical trial enrollment by reducing key barriers to large-scale screening for biomarker-determined eligibility^1,2^. High performing plasma assays of pTau217 and Aβ42/Aβ40 that perform comparably to gold standard CSF Aβ42/Aβ40 and Aβ PET imaging for detecting abnormal amyloid-β are now commercially available^3^, including those based on mass spectrometry^2^ (e.g., C_2_N Diagnostics) as well as immunoassays from Fujirebio Diagnostics on the fully-automated Lumipulse platform as well as from Quanterix using Simoa^4^. Blood-based biomarkers are also on a trajectory to transform clinical care by supporting more accurate diagnosis and determining eligibility for therapeautics^5,6^. In a recent landmark decision, the Lumipulse pTau217/Aβ42 ratio from Fujirebio was the first blood-based biomarker to be cleared by the FDA to support AD diagnosis in clinically impaired individuals, setting the stage for future approval of other high performing plasma assays. Given that Aβ-positive (A+) clinically unimpaired (CU) individuals are at increased risk for cognitive decline^7^ and it is likely that interventions are most likely to be successful when implemented early, it is critical that blood-based biomarkers are sensitive to early and subtle AD pathology in older CU populations.

Critically, Aβ and tau biomarkers offer only a partial picture of the complex underlying pathophysiology of AD and risk of clinical progression. For example, the presence of co-pathology such as vascular injury, ⍺-synuclein, and TDP-43 is common and impacts clinical trajectories^8^. Moreover, mounting evidence points to the relevance of additional biological processes that are not specific to AD, with markers of neuronal injury (e.g., neurofilament light chain, NfL), neuroinflammation (e.g., glial fibrillary acidic protein, GFAP), and synaptic integrity (SNAP-25, NPTX2) providing unique information regarding disease stage and risk for clinical progression^9–12^. Given the relevance of a growing number of plasma biomarkers, the need for multi-analyte blood-based biomarker panels that can measure multiple proteins simultaneously (i.e., multiplexing) is increasingly evident. For example, targeted multi-analyte panels could enable enhanced biomarker staging and theragnostic evaluation of possible interventions, both in clinical settings for treatment as well as in research settings to facilitate trial enrollment, setting the stage for personalized precision medicine. Until recently, there have not been any methods that have combined multiplex capabilities with AD biomarker specificity. For instance, high throughout multiplex proteomics platforms such as SomaScan and Olink have supported the discovery of novel proteins related to and independent of Aβ and tau that are linked to disease progression and cognitive impairment^10,13^, however neither reliably measures individual AD-specific markers (e.g., Aβ42, pTau217, pTau231, pTau181, etc.).

NULISA (Nucleic acid-Linked Immuno-Sandwich Assay)^14^ is a recently developed multiplex plasma proteomics platform that can simultaneously measure 124 targets with attomolar sensitivity and broad dynamic range, including core AD biomarkers as well as proteins related to inflammation, synaptic integrity, and neuronal injury. Initial studies using this platform with predominantly CU cohorts suggest that it performs well in measuring core AD biomarkers (pTau217, pTau231, pTau181, Aβ42) as well as established markers of neuronal injury (NfL) and inflammation (GFAP) relative to Simoa immunoassays^15,16^. Moreover, recent work has demonstrated comparable performance of NULISAseq plasma pTau217 and Simoa plasma pTau217 for detection of Aβ PET positivity in predominantly CU cohorts^15,17^ and across the AD clinical spectrum^18^. Thus, NULISA may be well-suited to support measurement of known AD biomarkers together with measurement of related processes that could enable the development and validation of multi-analyte biomarker panels for use across both research and clinical settings.

While these initial studies suggest NULISA holds significant promise, there remains a need to validate NULISAseq assays against additional high-performing plasma immunoassays in independent CU cohorts, determine their sensitivity to gold standard CSF biomarkers of Aβ and tau pathology among CU, and identify additional biological pathways relevant to early disease processes and subtle memory impairment in preclinical AD. The current study addresses this gap by conducting a head-to-head comparison of NULISAseq and Lumipulse plasma immunoassays in CU older adults enrolled in the Stanford Aging and Memory Study. The first aim of the current study was to evaluate performance of the NULISAseq CNS panel for measurement of core AD biomarkers pTau217, pTau181, Aβ42, Aβ42, Aβ42/Aβ40, pTau217/Aβ42, as well as NfL and GFAP, through comparison with corresponding Lumipulse assays. The second aim was to compare the discriminative accuracy of NULISAseq and Lumipulse plasma assays for detecting CSF-defined Aβ-positivity among CU. The third aim was to identify novel plasma markers associated with AD risk factors, including age, female sex, and *APOE-*ε4, as well as plasma markers differentially expressed in relation to gold standard biomarkers CSF Aβ42/Aβ40, CSF pTau181, temporal cortex tau accumulation, as well as memory function among CU.

## Method

### Participants

This study included 193 participants enrolled in the Stanford Aging and Memory Study (SAMS)^19,20^. Plasma samples were collected at baseline (*n*=162), or during a follow-up visit (*n*=31) conducted ∼3.5 (*n*=27) or ∼7 (*n*=4) years after baseline enrollment. Clinical diagnosis was determined at a clinical consensus meeting by a panel of neurologists and neuropsychologists based on a comprehensive neuropsychological battery. SAMS eligibility for baseline enrollment required a consensus diagnosis of CU, a Clinical Dementia Rating (CDR) score^18^ of 0 and performance within 1.5 standard deviations of demographically-adjusted means of standardized neuropsychological assessments. Of participants that were followed over time (*n*=31), 29 participants had a CDR of 0 at their most recent visit, one participant had a CDR of 0.5 with a CU diagnosis, and one participant was missing CDR and clinical diagnosis. The individual missing CDR was within 1.5 standard deviations of demographically-adjusted means across standardized neuropsychological assessments at follow-up. Other eligibility criteria included normal or corrected to normal vision and hearing, right-handedness, native English speaking, and no history of neurological or psychiatric disease. Study protocols were approved by the Institutional Review Board of Stanford University and written informed consent was obtained from each study participant.

### Plasma collection and analysis

EDTA plasma was collected by venipuncture, centrifuged for 10 minutes at 2000 x g, aliquoted in polypropylene tubes, and stored at -80°C until measurement. 193 samples from unique participants were available for NULISAseq analyses. Of the 193 unique participants with NULISA data, Lumipulse plasma pTau217, GFAP, and NfL was available in all 193 participants, plasma Aβ42 and Aβ40 were available in 188 participants, and plasma pTau181 was available in 186 participants.

### NULISAseq CNS disease panel

Plasma samples were analyzed at the Stanford University Human Immune Monitoring Center using the NULISAseq CNS kit on 7/31/2024 (Alamar Part# 800104, Lot# 05202024). Samples were thawed at room temperature. Thawing period does not exceed 3 hours, centrifuged for 1 min at room temperature at 1000 x g if there are no bubbles in the sample plate; otherwise, centrifuged for 5 min, RT at 1258 x g to remove bubbles, and loaded on an Alamar ARGO™ system. The following steps were automated on the instrument: Following incubation with capture and detection antibodies conjugated with partially double-stranded DNA containing target-specific barcodes, solutions underwent magnetic bead-based capture, wash, release, recapture, and second round of wash processes to identify bound immunocomplexes. DNA reporter molecules were generated through ligation and quantified by Next-Generation Sequencing (NGS) using an Element AVITI sequencer (Element Biosciences).

The CNS panel included 124 targets. NULISA Protein Quantification (NPQ) units were used in subsequent analyses. NPQ units are target counts that were normalized in two steps and then log2-transformed. For normalization, counts were divided by internal control counts within each sample well and then subsequently divided by target-specific medians of sample replicates on each plate. The average coefficient of variation (CV) of the triplicate sample control measures across the 124 targets was 6.0%.

### Lumipulse plasma analysis

Lumipulse plasma analyses were completed by the Stanford ADRC Biomarker Core and included pTau181 (5/2021), Aβ42 and Aβ40 (5/2023), and pTau217, GFAP, and NfL (12/2024). For each analysis, plasma samples were thawed on wet ice, centrifuged for 5 min at 4°C at 1,000 x g, and loaded on a Lumipulse *G* 1200 instrument (Fujirebio US, Malvern, PA) as previously described^21^. Given the fully-automated nature of the Lumipulse instrument, duplicate measures are expected to yield near-identical results. Accordingly, samples are typically measured in singlicate within each batch.

### CSF collection and analysis

CSF data were included for 132 participants with a CSF draw within 1 year of their plasma draw. CSF was collected by lumbar puncture and stored in polypropylene tubes. CSF samples were centrifuged, aliquoted, and stored at -80°C until measurement. CSF AD biomarkers Aβ42, Aβ40, and pTau181 were measured using the fully-automated Lumipulse *G* 1200 instrument as previously described^22^. CSF Aβ and Tau positivity cut-offs were defined using the SAMS baseline sample^20^ (*n*=153, **Supplementary** Figure 1). CSF Aβ positivity was defined as Aβ42/Aβ40 < 0.0752, corresponding to the 0.5 probability of belonging to the positive distribution using Gaussian mixture-modelling^20^. CSF Tau positivity was defined as pTau181 > 50.77, corresponding to 2SD above the mean of the Aβ-negative group.

### Amyloid and Tau PET imaging

Aβ PET measured using 18F-florbetaben was used to determine Aβ status for 25 participants with a PET scan within 2 years of their plasma measurement or a positive scan that occurred prior to plasma measurement. Tau PET measured using 18F-PI2620 was included for 63 participants with a Tau PET scan collected within 2 years of baseline plasma draw (mean (SD) delay = 0.56 (0.618) months). Aβ and Tau PET scanning was completed at the Richard M. Lucas Center for Imaging at Stanford University using a PET/MRI scanner (Signa 3T, GE Healthcare). For Aβ PET, emission data were collected between 90-110 minutes. For Tau PET, emission data were collected between either 45-75 minutes or 60-90 minutes (60-90-minute data was interpolated to produce 45–75-minute images^34^). PET images were reconstructed using TOF optimized subset expectation maximization (TOF-OSEM) with 3 iterations, 28 subsets, and 2.78 × 1.17 × 1.17 mm voxel size. Corrections were applied for detector deadtime, scatter, randoms, detector normalization, and radioisotope decay. MR attenuation correction was performed with ZTE MR imaging^22^. PET data were reconstructed into 5-minute frames and these frames were realigned and summed. Native space FreeSurfer labels defined on each participant’s T1-weighted MPRAGE structural MRI were used to extract intensity values from the co-registered summed PET data and used to create standardized uptake value ratios (SUVRs). Aβ PET SUVRs were created for a global cortical ROI (an average across frontal, parietal, lateral temporal, and cingulate) using a whole cerebellum reference region^23^ and translated to centiloids^24^. A threshold of >25 CL was used for Aβ positivity^4^. Tau PET SUVR was computed using the inferior cerebellum as a reference region for a temporal meta-ROI (entorhinal cortex, amygdala, parahippocampal, fusiform, inferior temporal, and middle temporal gyri)^23^.

### Memory composite score

A delayed recall composite score was computed from the delayed recall scores from 1) the Logical Memory subtest of the Wechsler Memory Scale; 2) the Hopkins Verbal Learning Test-Revised; and 3) the Brief Visuospatial Memory Test– Revised. Composite scores were computed by first z-scoring individual subtest scores using the full SAMS baseline CU sample (*n* = 212) as reference and then averaging^20^.

### Statistical analysis

All statistical analyses were conducted using R software version 4.3.0. The strength and direction of associations between plasma proteins measured using NULISAseq and Lumipulse assays were assessed using both Pearson and Spearman Rank correlation. Plots also show a regression fit and 95%CI. Receiver operating characteristic (ROC) curve analysis was performed to summarize the ability of individual proteins and protein ratios to differentiate CSF Aβ status. DeLong’s test was used to compare ROC AUC (area under the curve) across models. Performance of individual assays was also compared to a basic model including age, sex, and *APOE*-ε4 carriage. The Youden method was used to determine the optimal plasma cut-off to maximize sensitivity and specificity in discriminating Lumipulse CSF A+ from A-individuals.

Volcano plots were used to display univariate associations between variables of interest and NULISAseq CNS targets (N=124). Linear regression models were used to evaluate associations between NULISAseq CNS targets and (1) age, sex, and *APOE*-ε4, (2) CSF Aβ42/Aβ40 and pTau181, adjusted for age and sex, and (3) memory (delayed recall composite score), adjusted for age, sex, and education. Volcano plots display beta coefficients and log10 *p*-values. To evaluate differential expression of NULISAseq CNS targets by CSF biomarker-defined groups of interest (A+ vs A-; T+ vs T-; A-T-vs A+T-; A+T-vs A+T+), fold change differences between groups were calculated as 2^(difference^ ^in^ ^NPQ)^. Absolute log2 fold change differences of at least 0.2 was used as a criterion to determine effects of interest. Wilcoxon rank-sum tests were used to evaluate the significance of associations between fold change differences in NPQs and biomarker-defined groups without adjustment for covariates. False discovery rates (FDRs) were calculated using the Benjamini and Hochberg procedure^24^. A *p-*value < 0.05 and FDR <5% was considered statistically significant, whereas a *p*-value of < 0.05 and FDR >5% was considered marginally significant.

## Results

### Cohort characteristics

This study included data from 193 participants from the SAMS cohort (mean age 69.9 (6.35) years, 56% women, 86% non-Hispanic White; see **Table 1** for demographic characteristics). Of participants with known Aβ status (*n*=158), 25.9% were Aβ+ by CSF (N=37) or Aβ-PET (N=4). Of participants with available CSF (*n=*132), 15.9% (*n*=21) were Tau+.

**Table 1:** Sample Demographics.

|  | **SAMS CU (n = 193)** |
| --- | --- |
| **Age, mean (SD) y** | 69.9 (6.35) |
| **Sex, n female (% female)** | 108 (56.0%) |
| **Education, mean (SD) y** | 16.8 (2.04) |
| **APOE genotype** |  |
| $\epsilon 2/\epsilon 3$ | 16 (8.3%) |
| $\epsilon 2/\epsilon 4$ | 3 (1.6%) |
| $\epsilon 3/\epsilon 3$ | 131 (67.9%) |
| $\epsilon 3/\epsilon 4$ | 39 (20.2%) |
| $\epsilon 4/\epsilon 4$ | 4 (2.1%) |
| **Amyloid Status<sup>a</sup> (n Aβ+ / n total)** | 41/158 (25.9%) |
| **CSF Aβ42/Aβ40, mean (SD)** | 0.0877 (0.0232) |
| **CSF pTau181, mean (SD)** | 39.5 (22.1) |
| **Race** |  |
| Asian | 15 (7.8%) |
| Black or African American | 4 (2.1%) |
| White | 166 (86.0%) |
| More Than One Race | 5 (2.6%) |
| Other | 1 (0.5%) |
| Unknown/Not Reported | 2 (1.0%) |
| **Ethnicity** |  |
| Hispanic or Latino/a | 9 (4.7%) |
| NOT Hispanic or Latino/a | 184 (95.3%) |

<sup>a</sup> Aβ status determined by CSF (n=133, 37 Aβ +) or Aβ PET (n=25, 4 Aβ +)

### Head-to-head comparison of NULISAseq and Lumipulse plasma assays

We first examined the correlation between NULISAseq measurements and Lumipulse measurements of core AD-related plasma proteins, including Aβ42, Aβ40, pTau217, pTau181, GFAP, and NfL. We also examined plasma Aβ42/Aβ40 and pTau217/Aβ42 ratios across platforms. All pairwise comparisons exhibited moderate to high correlations, with Spearman correlation coefficients ranging from 0.50-0.93 and Pearson correlation coefficients ranging from 0.43-0.94 (**Fig. 1**). The strongest correlations were observed for GFAP (*rho =* 0.93; *r =* 0.93) and NfL (*rho =* 0.92; *r =* 0.94). The weakest association was observed for the Aβ42/Aβ40 ratio (*rho =* 0.43, *r* = 0.50). The correspondence across platforms between pTau measures was moderately high (*rho* values ranging between 0.60 and 0.72; *r* values between 0.68 and 0.71).

**Fig. 1.**
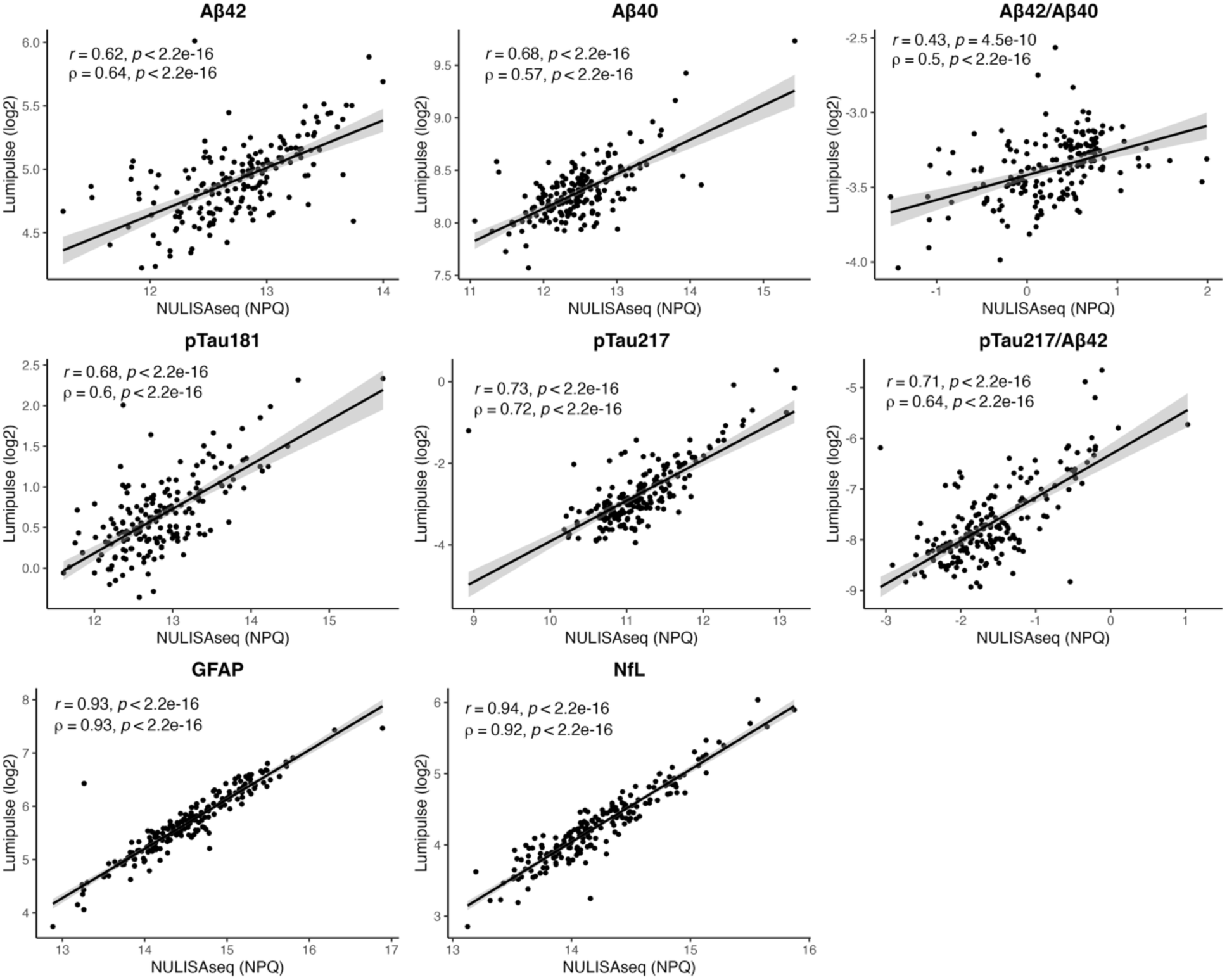
Correlations between NULISAseq and Lumipulse plasma assays. Scatterplots illustrate the distributions and correlations of proteins and protein ratios measured using the NULISAseq and Lumipulse platforms. NULISAseq proteins are in NPQ units and Lumipulse proteins measured in pg/mL were log2 transformed for comparison to NULISAseq. Pearson (*r*) and Spearman (*ρ*) correlation coefficients and *p-*values are shown. Scatterplots also show regression lines and 95% confidence intervals (shaded area). NPQ: NULISA protein quantification unit.

We next conducted ROC curve analyses to evaluate the performance of plasma protein ratios Aβ42/Aβ40 and pTau217/Aβ42 as well as individual proteins pTau217, pTau181, GFAP, and NfL in discriminating CSF Aβ positivity in CU (**Fig. 2A**). Across platforms, plasma pTau217/Aβ42 exhibited the highest performance in discriminating CSF Aβ positivity in CU, with an AUC of 0.940 (95% CI: 0.885 - 0.995) for NULISAseq and an AUC of 0.907 (95% CI: 0.849 - 0.966) for Lumipulse. The performances of these two assays did not significantly differ (DeLong’s, Z = 1.56, *p* = 0.119). This ratio significantly outperformed the covariate only-model across NULISAseq (Z = 3.89, *p* < 0.001) and Lumipulse platforms (Z = 3.89, *p* = 0.002).

**Figure 2:**
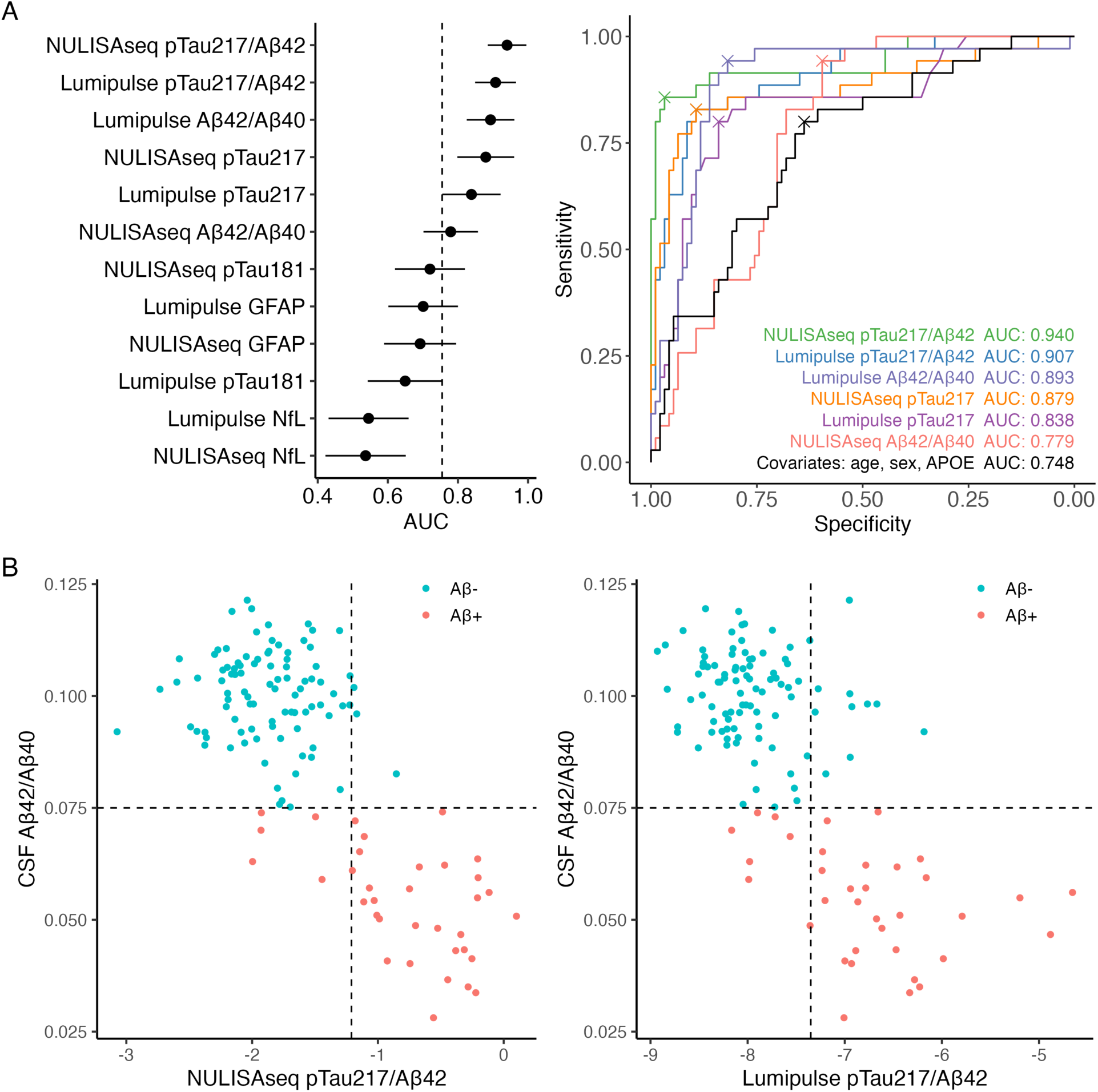
Performance of NULISAseq and Lumipulse plasma assays in detecting CSF Aβ positivity. **A** Forest plot depicting AUC and 95% CI for individual assays and biomarker ratios in detecting CSF Aβ positivity (left). The dotted line indicates the AUC of a covariate-only model including age, sex, and *APOE-*ε4. Corresponding ROC curves for the top seven performing models across platforms and the covariate-only model (right). **B** Concordance between CSF Aβ42/Aβ40 and plasma pTau217/Aβ42 measured using NULISAseq and Lumipulse. Dotted lines indicate the Aβ positivity cut-offs for each modality, giving rise to four participant profiles. AUC: Area under the curve.

Similarly, plasma pTau217 alone exhibited strong performance across platforms, with a NULISAseq AUC of 0.879 (95% CI: 0.798, 0.960) that significantly outperformed the covariate-only model (*Z* = 2.62, *p* = 0.009) and Lumipulse AUC of 0.838 (95% CI: 0.755, 0.922) that marginally outperformed the covariate-only model (*Z* = 1.66, *p* = 0.097). The performance of these two assays did not significantly differ (Z = 1.66, *p* = 0.097). In contrast, Lumipulse Aβ42/Aβ40 (AUC: 0.893, 95% CI: 0.825, 0.961) significantly outperformed NULISAseq Aβ42/Aβ40 (AUC: 0.779, 95% CI: 0.702, 0.857; Z = 2.66, *P* = 0.008), with NULISAseq Aβ42/Aβ40 not significantly outperforming a covariate-only model (*Z* = 0.55, *p* = 0.582). Models including pTau181 or GFAP alone did not perform significantly better than a covariate-only model (all *p* > 0.12), while models with NfL alone performed significantly worse than the covariate-only model across platforms (NULISAseq: *Z* = 3.89, *p* < .001; Lumipulse: *Z* = 3.70, *p <* 0.001). Performance of pTau181, GFAP, and NfL did not differ across platforms (all *p* > 0.10).

Selecting plasma pTau217/Aβ42 as the top performing assay for predicting CSF Aβ positivity, we derived a threshold to maximize sensitivity and specificity using the Youden method. We observed strong agreement across modalities using the NULISAseq pTau217/Aβ42-defined Aβ threshold (**Fig. 2B**), such that 93.8% of participants (*n*=121) had concordant plasma and CSF profiles and 6.2% (*n*=8) had discordant profiles. We also observed strong agreement across modalities with the Lumipulse pTau217/Aβ42-derived Aβ threshold (**Fig. 2B**), with 87.6% of participants (*n=*113) exhibiting concordant plasma and CSF profiles, however twice as many participants (12.4%, *n*=16) exhibited discordant profiles compared to that observed with the NULISAseq ratio.

### NULISAseq plasma CNS panel targets associate with age, sex, and *APOE-*ε4

We next examined associations between NULISAseq CNS plasma proteins and common AD risk factors, age, sex, and *APOE-*ε4 carriage (**Fig. 3**). A total of 40 targets (33% of CNS proteins) exhibited significant associations with age. These included proteins implicated in neurodegeneration and axonal injury, including NFL and neurofilament heavy polypeptides (NEFH), fatty acid-binding protein 3 (FABP3), and cystatin C (CST3), multiple pTau (pTau231, pTau181) and Aβ (Aβ42, Aβ40, Aβ38) isoforms. Also associated with age were proteins relating to microglial activation and inflammation, including triggering receptor expressed on myeloid cells 1 (TREM1) and 2 (TREM2), GFAP, serum amyloid A1 (SAA1), chitinase-3-like protein 1 (CHI3L1/YKL-40), along with several cytokines (IL6, IL33, IL16) and chemokines (CCL4, CCL3, CCL11, CXCL10). While most proteins were upregulated with age, glial cell line-derived neurotrophic factor (GDNF) and corticotropin-releasing hormone (CRH) were downregulated with age.

**Figure 3.**
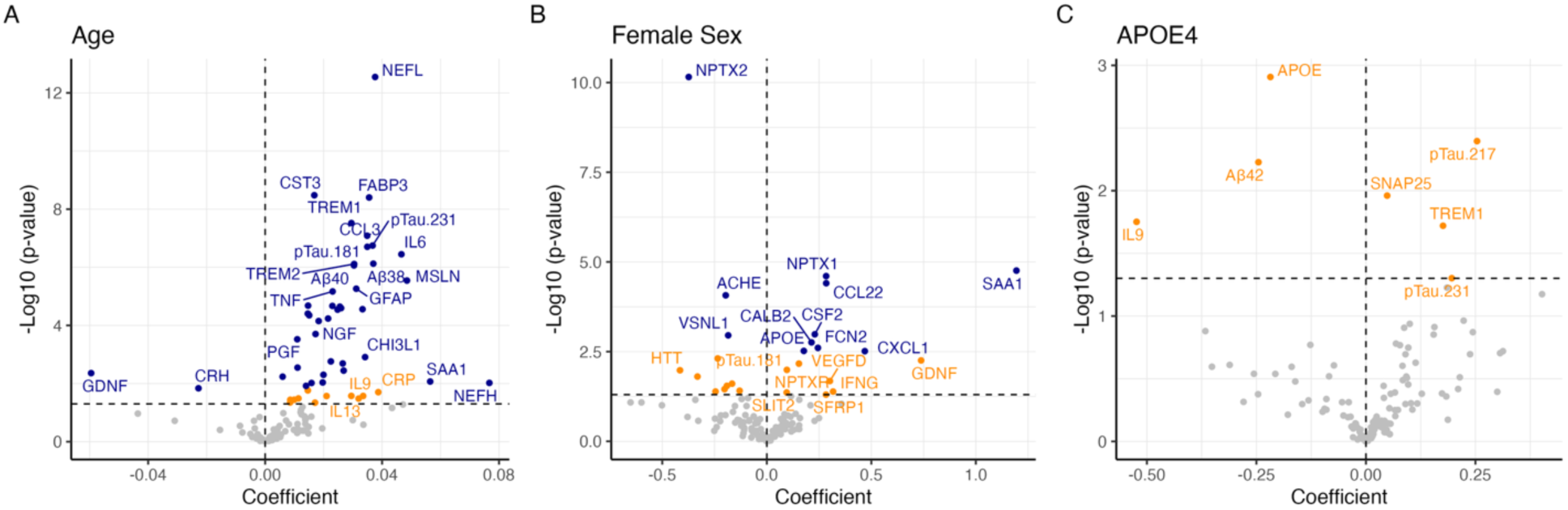
NULISAseq plasma protein associations with age, sex, and *APOE-*ε4. **A** Volcano plots show regression coefficients and -log10 (*p-*value) from models assessing associations between NULISAseq targets and age, **B** female sex, and **C** *APOE-*ε4 carriage. Significant targets (*p <* 0.05 and FDR <5%) are shown in blue, marginally significant targets (*p <* 0.05 and FDR >5%) are shown in orange, and non-significant targets (*p >* 0.05) are shown in gray.

Sex differences were observed across 10 targets (8% of CNS proteins). Proteins most elevated in females included several targets involved in the innate immune response, including SAA1, ficolin-2 (FCN2), the cytokine colony-stimulating factor 2 (CSF2), chemokines CCL22 and CXCL1, as well as neuronal pentraxin 1 (NPTX1) and calretinin (CALB2), a calcium binding protein that modulates neuronal excitability. Proteins most elevated in males compared to females included neuronal pentraxin 2 (NPTX2), acetylcholinesterase (ACHE), and visinin-like protein 1 (VSNL1). *APOE-*ε4 carriage was linked to marginally significant elevations in pTau217, SNAP-25, TREM1, and pTau231 and lower levels of Aβ42 and IL9.

### CSF amyloid and tau pathology associate with NULISAseq plasma CNS panel targets

We next examined associations between NULISAseq CNS plasma targets and continuous CSF Aβ42/Aβ40 and CSF pTau181, controlling for age and sex. As expected, abnormal CSF Aβ42/Aβ40 was associated with higher levels of plasma pTau217, pTau231, pTau181, and GFAP as well as lower Aβ42 (**Fig. 4A**). Elevated CSF pTau181 was associated with higher levels of plasma pTau217, pTau231, pTau181, GFAP, as well as pro-inflammatory chemokine CXCL8 (IL8) (**Fig. 4B**).

**Fig 4.**
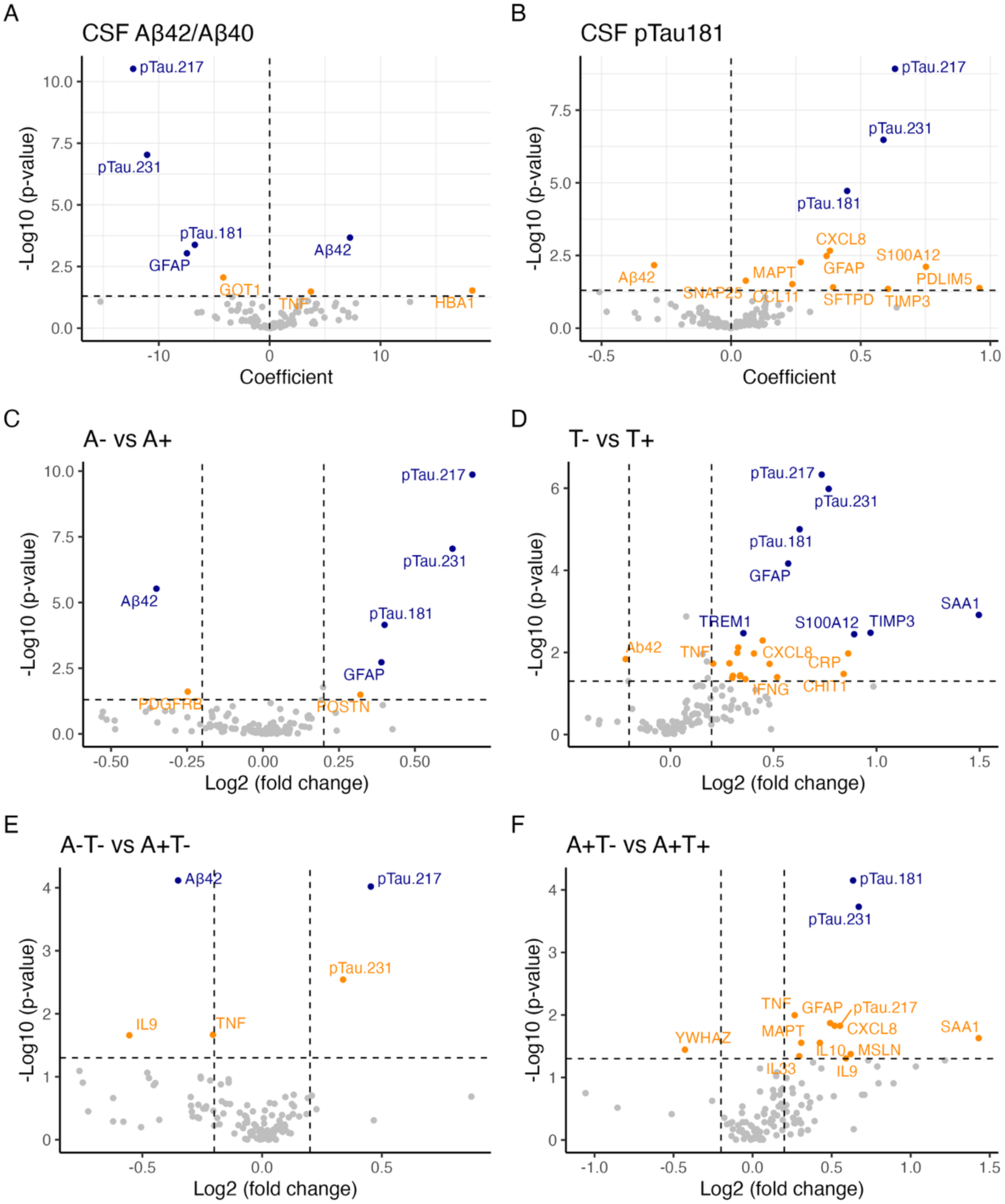
Association of NULISAseq plasma targets with Aβ and tau pathology. **A** Volcano plots illustrating NULISAseq plasma targets associated with CSF Aβ42/Aβ40 and **B** CSF pTau181, with age and sex as covariates. **C** Fold change differences in NULISAseq plasma targets between A- (*n*=96) vs A+ (*n*=36) groups, **D** T- (*n*=111) vs T+ (*n*=21) groups, **E** A-T- (*n*=91) vs A+T- (*n*=20) groups, and **F** A+T- (*n*=20) vs A+T+ (*n*=16) groups. Absolute fold change differences greater than 0.02 were considered effects of interest. Significant targets (*p <* 0.05 and FDR <5%) are shown in blue, marginally significant targets (*p <* 0.05 and FDR >5%) are shown in orange, and non-significant targets (*p* > 0.05) are shown in gray.

To further probe protein targets that varied specifically with abnormal CSF Aβ (A+) and tau (T+) positivity among CU older adults, we examined fold-change differences in NULISAseq targets as a function of CSF-defined A and T status groups. First, in comparing A- and A+ groups only (**Fig. 4C**), we observed differential expression of the identical set of proteins that were associated with continuous CSF Aβ42/Aβ40, including plasma pTau217, pTau231, pTau181, and GFAP, and Aβ42. This underscores the strong link between pTau proteins and astrocytosis and the development of CNS amyloidosis. Next, comparing T- and T+ groups (**Fig. 4D**), we again observed upregulation of plasma pTau217, pTau231, pTau181, and GFAP, together with an additional four targets linked to immune signaling and inflammation, including TREM1, tissue inhibitor of metalloproteinases-3 (TIMP3), SAA1, and S100 calcium-binding protein A12 (S100A12) a proinflammatory calcium binding protein primarily expressed in immune cells.

To identify differentially expressed proteins linked to early Aβ, prior to elevations in CSF pTau181, we compared fold-change differences between individuals negative for both Aβ and tau (A-T-, *n*=91) and individuals who are Aβ-positive and Tau-negative (A+T-, *n*=20). A+T-CU exhibited significant downregulation of plasma Aβ42 and upregulation of pTau217, with marginally significant upregulation of pTau231. This highlights the role of plasma pTau217 as a sensitive marker of early CNS amyloid accumulation. To identify proteins that become elevated when both Aβ and tau biomarkers are abnormal, likely indicating further progression along the AD continuum^13^, we compared the A+T-group to individuals who were Aβ-positive and Tau-positive (A+T+, *n*=16). This analysis revealed significant upregulation of plasma pTau181 and pTau231, with a larger set of proteins exhibiting marginally significant elevations, including pTau217, GFAP, microtubule-associated protein tau (MAPT), along with a contingent of inflammatory proteins tumor necrosis factor (TNF), IL9, IL10, IL33, and CXCL8. The increased presence of these inflammatory molecules indicates that the well-established tau-associated inflammatory processes are initiated early, as demonstrated in these CU individuals.

### Association of NULISAseq plasma targets with temporal cortex tau burden

Plasma pTau biomarkers tend to exhibit stronger associations with cerebral Aβ and become abnormal prior to abnormal tau accumulation, which represents a later stage of progression along the AD continuum^25–27^. We next examined associations between NULISAseq plasma proteins and regional tau burden in temporal cortex in participants with Tau PET data collected within 2 years of plasma (**Table 2**). As expected, we observed significant associations between temporal tau accumulation and plasma pTau217 (β=0.083, *p* = 0.001), pTau217/Aβ42 (β=0.070, *p* < 0.00), pTau231 (β=0.060, *p* = 0.011), and pTau181 (β=0.056, *p* = 0.021; **Fig. 5A**). The strength of the associations between temporal cortex tau and plasma pTau217 and pTau217/Aβ42 were comparable to those measured using the Lumipulse platform (**Supplementary** Fig. 2). When examining associations of temporal cortex tau burden with the NULISAseq CNS panel, we observed marginally significant associations with several proteins (**Fig. 5B**), including upregulation of plasma pTau217, pTau231, and pTau181, as well as enolase 2 (ENO2), an enzyme that plays a crucial role in neuronal glycolysis. Moreover, several proteins exhibited marginally significant negative associations with temporal tau SUVR, including Aβ42 and inflammatory proteins IL5, CCL22, CSF2, and interferon-gamma (IFNG).

**Fig 5.**
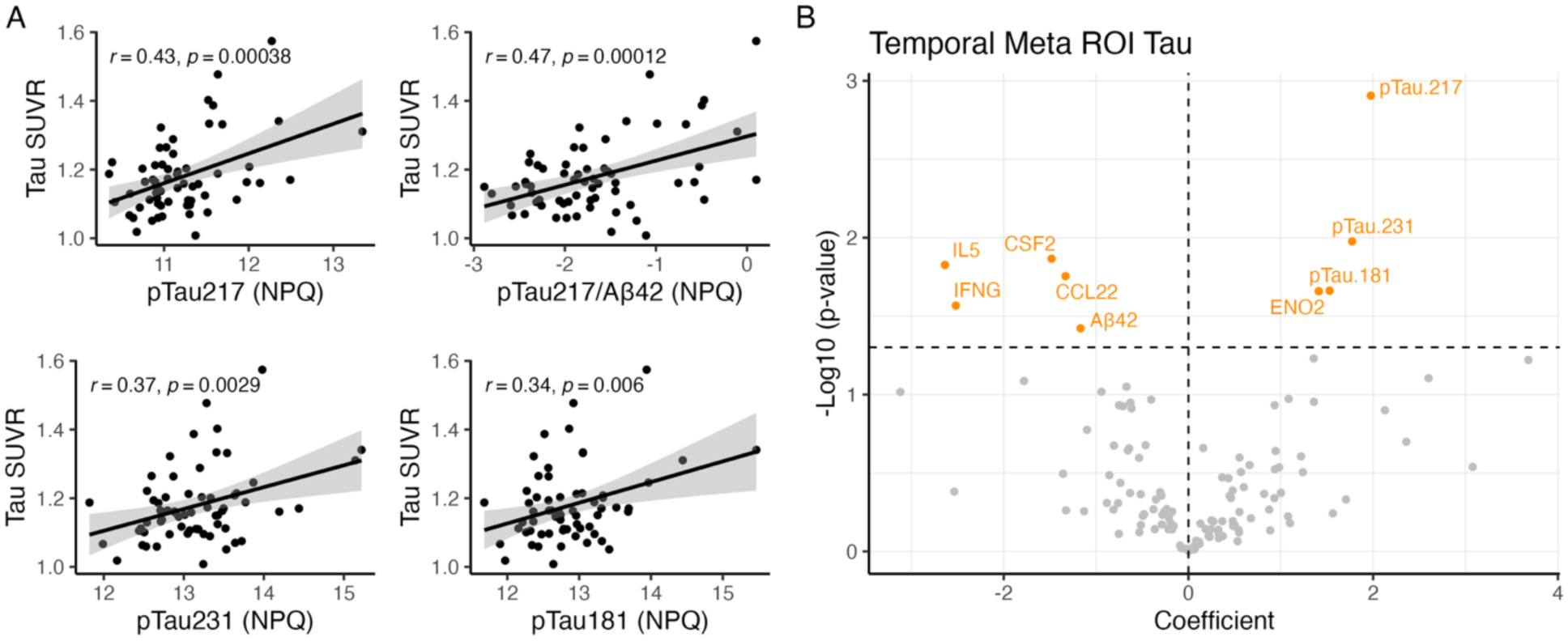
Association between NULISAseq plasma targets with temporal cortex tau accumulation. **A** Scatterplots depicting associations between temporal meta-ROI SUVR and NULISAseq plasma pTau isoforms in NPQ units. Scatterplots show regression lines, 95% confidence intervals (shaded area), and Pearson’s correlation coefficients. **B** Volcano plot showing associations between NULISAseq targets and temporal meta-ROI SUVR, adjusted for age and sex. Marginally significant targets (*p <* 0.05 and FDR >5%) are shown in orange, and non-significant targets (*p >* 0.05) are shown in gray. NPQ: NULISA protein quantification unit; SUVR: Standardized uptake value ratio.

These results once again highlight the key relationship between plasma inflammatory proteins detected by NULISAseq and tau neuropathology in CU.

### Association of NULISAseq plasma targets with memory function

Finally, we sought to identify NULISAseq plasma proteins that are sensitive to individual differences in memory function among CU older adults, adjusting for age, sex, and education. We first examined associations between memory and plasma pTau isoforms measured with the NULISA panel. We observed significant negative associations between memory composite score and plasma pTau217 (β= -0.21, *p* = 0.036), pTau231 (β= -0.19, *p* = 0.039), pTau181 (β=- 0.20, *p* = 0.034; **Supplementary** Fig. 3). The strength of the association between plasma pTau217 and memory was similar when measured with Lumipulse immunoassays (β= -0.19, *p* = 0.071; **Supplementary** Fig. 4) Analysis with the NULISAseq plasma CNS panel revealed seven plasma protein targets that were significantly negatively associated with memory composite score (**Fig. 6A, Supplementary** Fig. 5), including 14-3-3-protein zeta/delta (YWHAZ), ENO2, Peroxiredoxin 6 (PRDX6), arylsulfatase A (ARSA), YKL-40, IL8, and FCN2.

**Fig 6.**
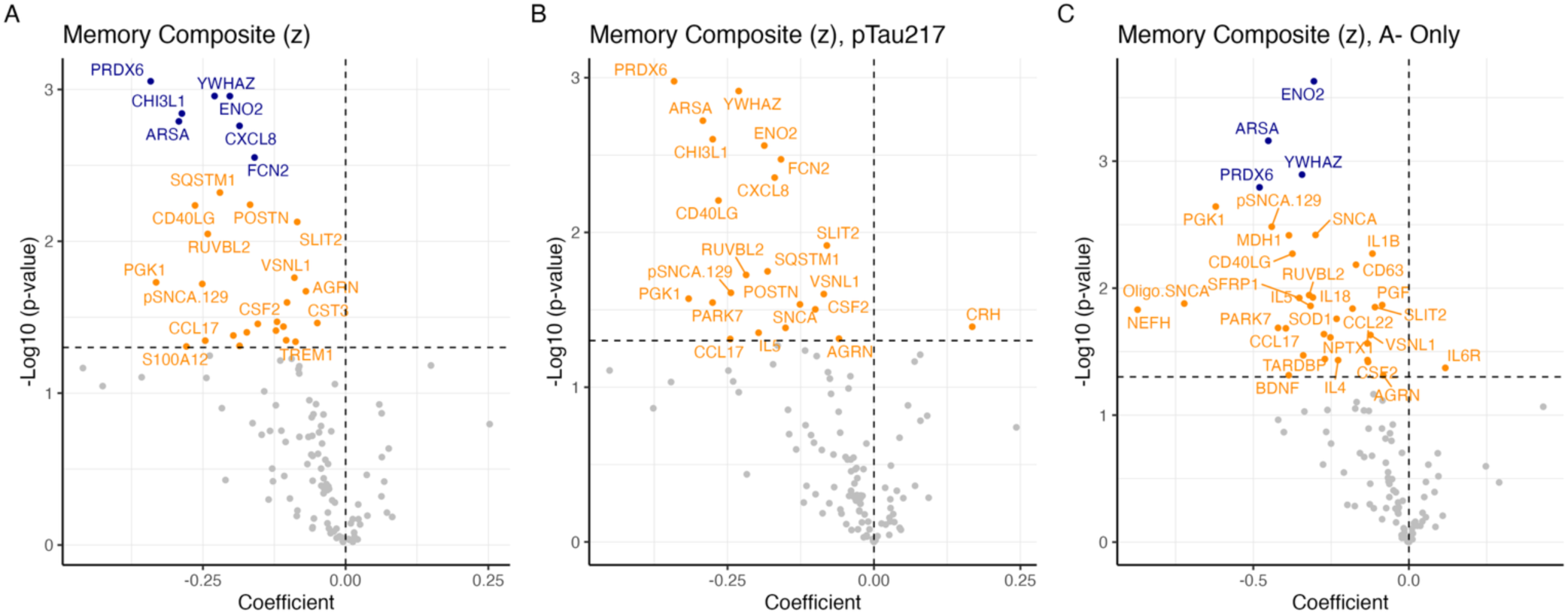
Association of NULISAseq plasma targets with memory. **A** Volcano plot illustrating NULISAseq targets associated with memory composite score, adjusted for age, sex, and education. **B** NULISAseq associations with memory, additionally controlling for pTau217. **C** NULISAseq associations with memory restricted to A-CU only. Significant targets (*p <* 0.05 and FDR <5%) are shown in blue, marginally significant targets (*p <* 0.05 and FDR >5%) are shown in orange, and non-significant targets (*p >* 0.05) are shown in gray.

To further examine if the identified targets explain unique variance in memory after accounting for core AD markers, we included NULISA plasma pTau217 as an additional covariate. The strength of the beta coefficients for each target remained effectively unchanged and FDR-corrected *p-*values were marginally significant (*p*_adj_ = 0.054; *p*_raw_ = 0.003; **Fig. 6B**), suggesting that the top associating memory-related proteins may be independent of plasma p-tau217 in this CU cohort. Similarly, when we restricted analysis to A-CU participants only (**Fig. 6C**), four of the initial seven plasma targets, ENO2, ARSA, YWHAZ, and PRDX6, remained significantly associated with memory, while plasma IL8 and YKL-40 exhibited marginally significant associations. Taken together, the set of NULISAseq CNS plasma proteins identified here implicate novel biological pathways that may become dysregulated in preclinical AD and contribute to early memory impairment.

## Discussion

Scalable, multi-analyte plasma proteomic panels that can accurately detect initial AD pathology and related pathophysiological processes relevant for disease risk among CU older adults are critical for early detection and prognosis. In this study, we evaluated the correspondence between plasma biomarkers measured using the NULISAseq CNS panel and the established fully-automated Lumipulse platform against gold standard CSF AD biomarkers in a CU cohort. Our results demonstrate strong alignment in plasma measurements across platforms, with moderate to high correlations across biomarkers of pTau, Aβ, NfL, and GFAP. NULISAseq pTau217/Aβ42 and pTau217 exhibited excellent diagnostic accuracy for detecting CSF-defined A+ status among CU that was equivalent to that demonstrated by corresponding Lumipulse measures. Moreover, we observed differential expression of several core AD-related biomarkers associated with CSF A+, with an expanded set of proteins related to inflammation and immune activation associated with CSF T+. Finally, we identified several NULISAseq targets that exhibited Aβ-independent associations with memory function, suggesting complementary biological pathways that may contribute to variability in cognitive function among CU older adults. Taken together, the present results provide robust evidence for sensitivity of NULISAseq assays to preclinical AD pathology and highlight the value of leveraging plasma multiplexing to characterize early abnormalities related to and independent of core AD biomarkers in the context of a healthy aging cohort.

The current results provide novel evidence for excellent correspondence between AD biomarkers measured using established Lumipulse immunoassays and the recently developed NULISA multiplex immunoassays among CU older adults. Specifically, we observed strong correlations between NULISAseq and Lumipulse plasma measurements of pTau217, pTau181, Aβ42, Aβ40, NfL, and GFAP, building on prior work demonstrating correspondence between NULISAseq and ALZpath Simoa immunoassays in predominantly CU cohorts^15–17^. In an important extension to past work, we also identified equivalent performance of plasma pTau217 and pTau217/Aβ42 ratios measured with Lumipulse and NULISAseq in detecting CSF A+ status within our CU cohort, with the pTau217/Aβ42 ratio exhibiting the highest accuracy across platforms [with a NULISAseq AUC of 0.940 (95% CI: 0.885, 0.995) and Lumipulse AUC of 0.907 (95% CI: 0.849 - 0.966)]. This observation is consistent with prior work demonstrating excellent performance of the pTau217/Aβ42 ratio measured using gold standard single-plex immunoassays in cohorts that span the clinical continuum^28,29^. Given existing research suggesting plasma ratios are less susceptible to measurement confounds related to kidney function and other comorbidities compared to individual analytes^28,30^, the pTau217/Aβ42 ratio may be particularly well suited for detecting A+ in real-world settings. Indeed, recent work comparing performance of the pTau217/Aβ42 ratio to pTau217 alone in clinical and community-based cohorts spanning the AD clinical continuum found that the ratio yielded fewer indeterminate cases that require follow-up biomarker testing^29^, a benefit that was more pronounced in the community-based cohort. The observed sensitivity of NULISA pTau217/Aβ42 to early Aβ accumulation in the current CU cohort further suggests that this ratio may be appropriate in the context of early detection and primary prevention trial enrollment.

Traditionally, measuring Aβ in plasma has been challenging compared to pTau isoforms. Here we found that plasma NULISA Aβ42/Aβ40 performed significantly worse (AUC 0.779) than plasma Lumipulse Aβ42/Aβ40 (AUC 0.893) in predicting CSF A+ status, and did not outperform a covariate model with age, sex, and *APOE-*ε4. This stands in contrast to plasma pTau217 and pTau217/Aβ42, which did not significantly differ across platforms and performed significantly better than the covariate model. While this finding regarding Aβ42/Aβ40 ratio requires replication in independent cohorts, it is consistent with prior work that found Aβ42 alone exhibited chance performance (AUCs 0.53-0.55) for detecting Aβ PET positivity across NULISAseq and Simoa platforms^15^. This observation suggests that the use of plasma Aβ42/Aβ40 as a “Core 1” biomarker in biological staging frameworks for AD^31^ is not appropriate for this version of the NULISA assay. While both Aβ42/Aβ40 and pTau217 are conceptualized as early changing Core 1 markers that are sufficient for determining whether an individual is on the AD trajectory^31^, these targets likely reflect different aspects of early changing pathophysiology. Indeed, prior work suggests combining measurements of Aβ42/Aβ40 and pTau217 can improve detection of abnormal Aβ pathology among CU^2^. Such findings are consistent with the strong performance of the pTau217/Aβ42 ratio for detecting Aβ+ in the current CU population as well as in past work^28,29^.

The current study also provided evidence for differential expression of NULISAseq Aβ42, pTau217, pTau231, and pTau181 plasma proteins in the context of abnormal gold standard Core 1 biomarkers reflecting initial AD pathological change, including CSF Aβ42/Aβ40 and CSF pTau181, as well as temporal cortex tau PET, a Core 2 biomarker index of further progression along the AD pathological continuum^31^. As in prior work using established single-plex immunoassays, associations of plasma Aβ42 and pTau isoforms were stronger with CSF biomarkers compared to tau accumulation in temporal cortex^25–27^, consistent with the classification of these plasma markers as Core 1 biomarkers. Interestingly, when participants were stratified by CSF-defined A/T groups to identify proteins linked to initial Aβ positivity the absence of abnormal CSF pTau181, we found that A+T-CU exhibited lower Aβ42 and elevated pTau217 relative to the biomarker negative group, whereas pTau181 and pTau231 were upregulated in the A+T+ group. This observation is consistent with the idea that Aβ42 and pTau217 are early changing biomarkers that reflect initial cerebral Aβ accumulation among CU, whereas pTau181 is more closely linked to downstream tau accumulation and neurodegeneration^32^.

There is growing appreciation of the role of maladaptive neuroinflammation in the pathophysiology and progression of AD, as indicated by the addition of the inflammation markers category in the revised AD biomarker framework^31^. Consistent with this idea, the present study identified multiple inflammatory biomarkers that were upregulated with abnormal AD pathology among CU, including GFAP, TREM1, and S100A12. Plasma GFAP, a measure of reactive astrogliosis, is thought to influence the relationship between Aβ accumulation and downstream tau phosphorylation and tangle accumulation^33^. The current observation of elevated GFAP with abnormal CSF Aβ42/Aβ40 complements prior work linking plasma GFAP to initial abnormal Aβ accumulation among CU^34–36^. TREM1, a pro-inflammatory myeloid cell receptor implicated in disruption of peripheral and brain immune cell metabolism and responses in aging, has been linked to clinical and neuropathological severity in AD^37^. In prior studies leveraging NULISA, S100A12, a pro-inflammatory calcium-binding protein, was differentially expressed among Tau PET-positive compared to Tau PET-negative CU^17^ and in AD compared to controls^38^ and may represent a novel biomarker of immune activation in AD^39^. The observation of elevations in these and other inflammatory markers in response to tau pathology among CU in the current study converge with a growing body of work implicating neuroinflammation in AD pathophysiology. Moreover, they provide additional evidence for early changes in multiple inflammatory markers in response to the early AD pathology among CU. These results motivate additional research leveraging multi-analyte plasma panels to characterize the impact of these early-changing inflammatory markers on progression to cognitive impairment.

Analyses examining associations between NULISAseq CNS targets and known AD risk factors such as age, sex, and *APOE-*ε4 identified a large set of proteins that may point to pathophysiological processes that underlie these risk profiles. For example, 40 CNS proteins were upregulated with age, including markers of neuronal injury (e.g., NfL, NEFH, FABP3), as well as pro-inflammatory cytokines and chemokines (e.g., IL6, CCL4) and regulators of microglial and astrocytic activation (e.g., TREM1, TREM2, and CHI3L1/YKL-40). Similarly, sex and *APOE-*ε4 were associated with differential expression of NPTX1, NPTX2 and SNAP25, markers of synaptic function that have been linked to risk and resilience to cognitive decline when measured in CSF^9,10^. While many of these proteins have been identified in prior work as markers of neurodegeneration, inflammation, and synaptic integrity that are relevant in aging and AD, they are typically only measured in the context of high throughput proteomic studies or highly targeted mass spectrometry studies. The ability to simultaneously measure these proteins alongside core AD biomarkers with multiplexing represents a significant benefit that can support greater understanding of the mechanisms underlying these AD risk factors and their role in cognitive decline and disease progression.

Prior work using gold standard biomarkers suggests that cross-sectional variance in memory function among CU is weakly linked to core AD proteins, but that much variance remains unexplained^20,40,41^. Consistent with existing evidence, we observed small but significant negative associations between NULISAseq pTau isoforms and memory in this CU cohort.

Interestingly, analyses examining associations between memory and the NULISAseq CNS panel revealed a set of proteins that are distinct from Aβ and tau biomarkers, including PRDX6, YWHAZ, CHI3L1/YKL-40, ARSA, ENO2, IL8, and FCN2. While replication in independent cohorts is needed, many of the targets observed here have been identified in prior work in relation to AD pathology or clinical phenotypes. For example, YKL-40, a marker of astrocytic activation, has been linked to tau pathology, neurodegeneration, and disease progression in AD when measured in CSF^35^. Higher levels of ENO2 in CSF, a protein related to neuronal energy metabolism, has been linked to greater tau fibrillar load and accumulation among CU^13^ and was upregulated with temporal cortex tau in the current study. Moreover, fold change increases in plasma PRDX6 and ARSA have been observed in MCI compared to CU using the NULISA CNS panel^17^, consistent with a possible role in cognitive impairment. Notably, the magnitude of the association of these targets with memory in the current study was similar in models controlling for NULISA pTau217, and in models including only Aβ negative participants, suggesting that these plasma proteins represent biological pathways that may become dysregulated among CU individuals that are relevant for cognitive function and are complementary to Aβ and tau. These results suggest that multi-analyte biomarker panels can support insights into pathophysiological processes that contribute to early cognitive impairment and may be leveraged in future work to improve prediction of risk for future cognitive decline in CU aging populations.

This study has limitations. Given modest rates of Aβ and tau positivity in this CU cohort, sample sizes for analyses examining fold change differences as a function of AD biomarker status are modest, and these findings require replication in larger cohorts. The current cohort is not representative of the broader population and future studies are needed to establish the performance of this NULISAseq technology in real-world settings. Current NULISAseq methods only allow relative quantification, which precludes the identification of a single cut off for AD-positivity that can be applied across different batches. As this new technology continues to develop, it will be important to provide absolute quantification for AD targets such as pTau217. Finally, these results are cross-sectional, and it will be important for future longitudinal studies to examine within-subject reliability and change in these markers over time, as well as determine the prognostic value of NULISAseq targets for predicting future cognitive decline and disease progression.

## Conclusions

Taken together, this study provides novel evidence for strong correspondence between core AD biomarkers measured across NULISAseq and Lumipulse plasma platforms and equivalent performance of plasma pTau217 and pTau217/Aβ42 for detecting CSF A+ among CU across platforms. These findings are significant, given the critical need for multi-analyte panels that can simultaneously measure known AD-relevant targets using a small blood volume without compromising sensitivity. Indeed, multiplexing enabled the identification of additional targets linked to inflammation, neuronal injury, and synaptic integrity that were associated with abnormal CSF Aβ and tau biomarkers, age, and memory function. These findings highlight the potential to leverage multiplexed proteomics to support rich phenotyping and characterization of mechanisms related to and independent of core AD biomarkers that contribute to early AD pathological change and cognitive impairment in older CU populations.

## Abbreviations

Aβ: Amyloid-beta
ACHE: acetylcholinesterase
AD: Alzheimer’s disease
AGRN: Agrin
APOE: apolipoprotein E
ARSA: arylsulfatase A
AUC: Area under Curve
CALB2: calretinin
CCL3 CC: chemokine ligand 3
CCL4 CC: chemokine ligand 4
CCL11 CC: chemokine ligand 11
CDR: Clinical dementia rating
CHI3L1: Chitinase-3 like-protein-1
CNS: Central nervous system
CRH: corticotropin-releasing hormone
CSF: Cerebrospinal fluid
CSF2: Granulocyte-macrophage colony-stimulating factor
CST3: cystatin C
CU: clinically unimpaired
CV: Coefficient of variation
CXCL10: chemokine ligand 10
CXCL8: chemokine ligand 8
ENO2: enolase 2
FABP3: fatty acid-binding protein 3
FDR: False discovery rate
GDNF: glial cell line-derived neurotrophic factor
GFAP: glial fibrillary acidic protein
IC: Internal control
IFNG: Interferon gamma
IL5: Interleukin 5
IL6: Interleukin 6
IL8: Interleukin 8
IL9: Interleukin 9
IL10: Interleukin 10
IL16: Interleukin 16
IL33: Interleukin 33
IQR: Interquartile range
IPC: Inter-plate control
LOD: Limit of detection
MAPT: microtubule-associated protein tau
MMSE: Mini-Mental State Examination
MRI: Magnetic resonance imaging
NEFH: Neurofilament heavy chain
NEFL: Neurofilament light chain
NPQ NULISA: protein quantification
NPTX1: Neuronal pentraxin-1
NPTX2: Neuronal pentraxin-2
NULISA: Nucleic acid-linked Immuno-Sandwich Assay
NULISAseq NULISA: with next-generation sequencing readout
PET: Positron emission tomography
PRDX6: Peroxiredoxin 6
p-tau: Phosphorylated tau
ROC: Receiver operating characteristic
ROI: Region of interest
S100A12 S100: calcium-binding protein A12
SD: Standard deviation
SAA1: serum amyloid A1
SAMS: Stanford Aging and Memory Study
Simoa: Single-molecule array
SNAP-25: Synaptosomal-associated protein, 25kDa
SUVR: Standardized uptake value ratio
TDP-43 TAR DNA: binding protein 43
TIMP3: Metalloproteinase inhibitor 3
TNF: tumor necrosis factor
TREM1: triggering receptor expressed on myeloid cells 1
TREM2: triggering receptor expressed on myeloid cells 2
YWHAZ: 14-3–3 Protein zeta/delta
VSNL1: visinin-like protein 1

## Supplementary Information

Additional file 1: Supplementary Table S1 and Supplementary Figures S1 to S5.

## Declarations

### Ethics approval and consent to participate

All participants provided written informed consent before participation in accordance with a protocol approved by the Stanford Institutional Review Board.

### Consent for publication

Not applicable.

### Availability of data and materials

Anonymized data are available upon request from qualified academic investigators.

## Author’s contributions

ANT, ECM, and ADW contributed to the study’s conception and design. IS, SW, DC, AA, KR, and TWC supported plasma data collection, processing, storage, and management. SJS supported CSF data collection. IS and SW supported Tau PET data collection and processing. TTN, HTM, ENW, and JM performed biochemical assays. ANT and KAC performed data analysis and produced figures. ANT wrote the manuscript. ECM, ADW, ENW, JW, and KAC were major contributors in writing the manuscript. All authors contributed to and approved the final version of the manuscript.

## Data Availability

Anonymized data are available upon request from qualified academic investigators.

## Acknowledgements

We thank all members of the Stanford Aging and Memory Study team and the SAMS participants for their valuable contributions to this research.

