## Supplemental Table 1, Supplemental Figures 1-5 for "Plasma proteomic signatures of preclinical Alzheimer’s disease biomarkers and memory in clinically unimpaired older adults"

**Supplementary Table 1: Demographic Characteristics of Tau PET sample**

| | **SAMS CU (n = 63)** |
|---|---|
| **Age, mean (SD) y** | 73.6 (6.51) |
| **Sex, n female (% female)** | 33 (52 %) |
| **Education, mean (SD) y** | 16.6 (2.16) |
| ***APOE* genotype** | |
| ε2/ε3 | 6 (9.5%) |
| ε2/ε4 | 1 (1.6%) |
| ε3/ε3 | 45 (71.4%) |
| ε3/ε4 | 9 (14.3%) |
| ε4/ε4 | 2 (3.2%) |
| **Amyloid Status (CSF)**<br>(n Aβ+ / n total) | 12 / 34 (35.3%) |
| **Race** | |
| Asian | 6 (9.5%) |
| Black or African American | 1 (1.6%) |
| White | 52 (82.5%) |
| More Than One Race | 2 (3.2%) |
| Unknown/Not Reported | 2 (3.2%) |
| **Ethnicity** | |
| Hispanic or Latino/a | 2 (3.2%) |
| NOT Hispanic or Latino/a | 61 (96.8%) |
| **Plasma, PET lag, mean (SD) y** | 0.56 (0.618) |

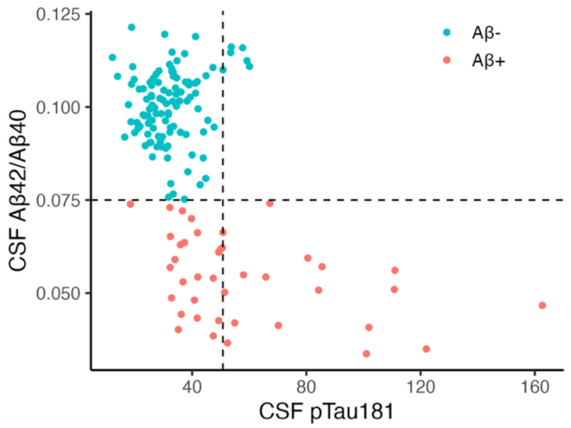


**Supplementary Figure 1.** CSF Aβ42/Aβ40 and pTau181 in SAMS CU. Scatterplot illustrating distributions and cut-offs for CSF-defined A and T groups in the SAMS baseline CU sample ($n$ = 153). Aβ positivity was defined as < 0.0752 and Tau positivity was defined as pTau181 > 50.77, corresponding to 2SD above the mean of the Aβ-negative group.

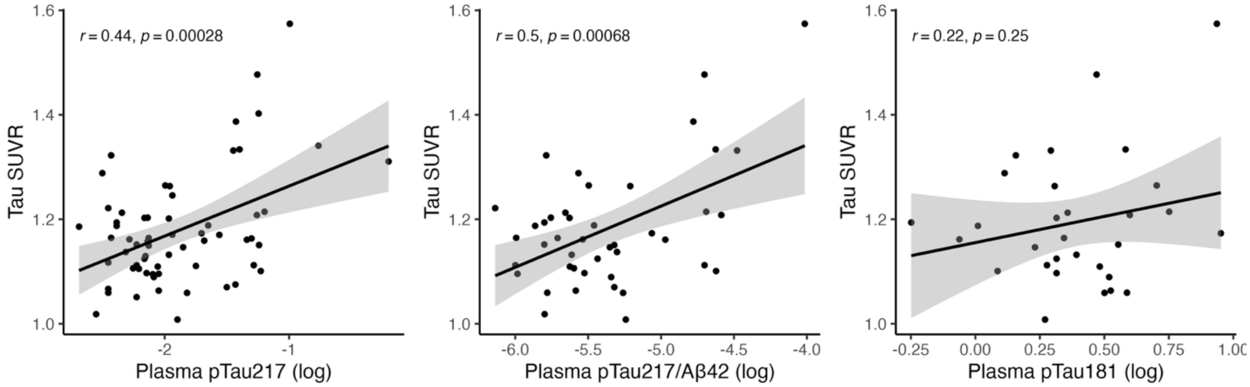


**Supplementary Figure 2.** Associations of Lumipulse plasma biomarkers with Tau PET SUVR in the meta-temporal cortex ROI. Scatterplots show regression lines, 95% confidence intervals (shaded area), and Pearson's correlation ($r$).

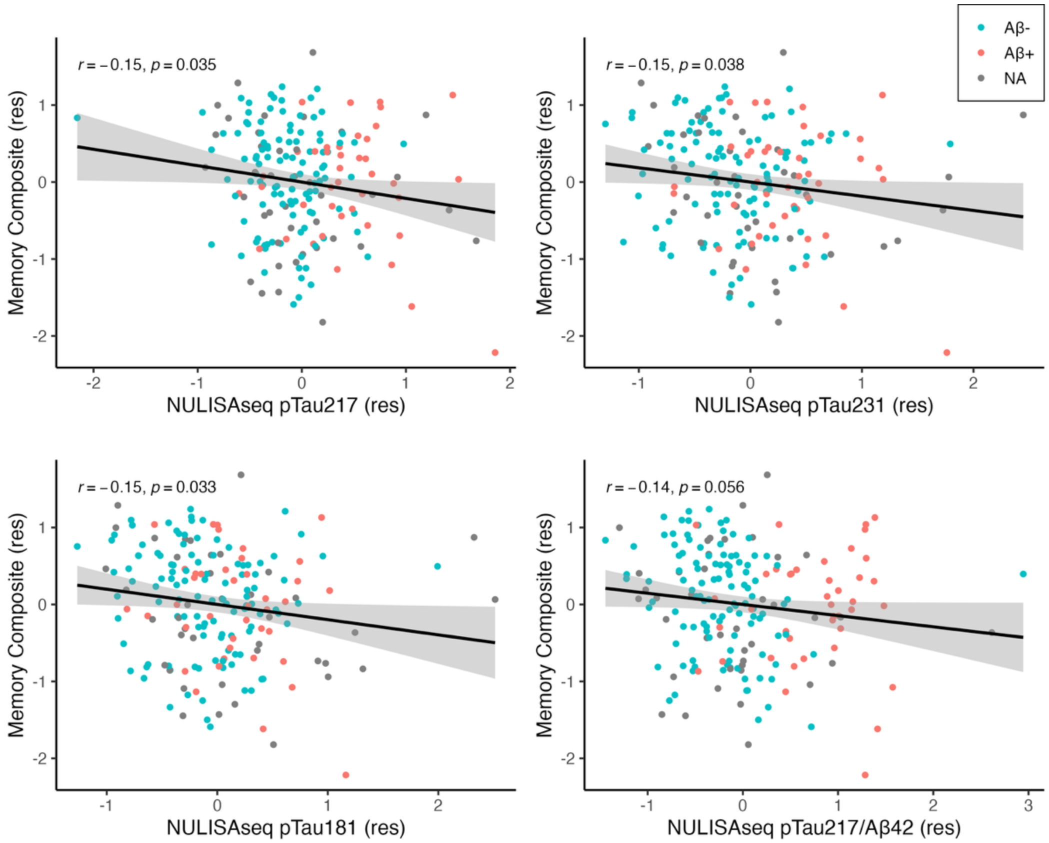


**Supplementary Figure 3.** Associations between NULISAseq plasma biomarkers and memory. Residuals controlling for age, sex, and education are plotted. Points are colored by Aβ status defined by CSF Aβ42/Aβ40 or Aβ PET. Scatterplots show regression lines, 95% confidence intervals (shaded area), and Pearson's correlation (*r*).

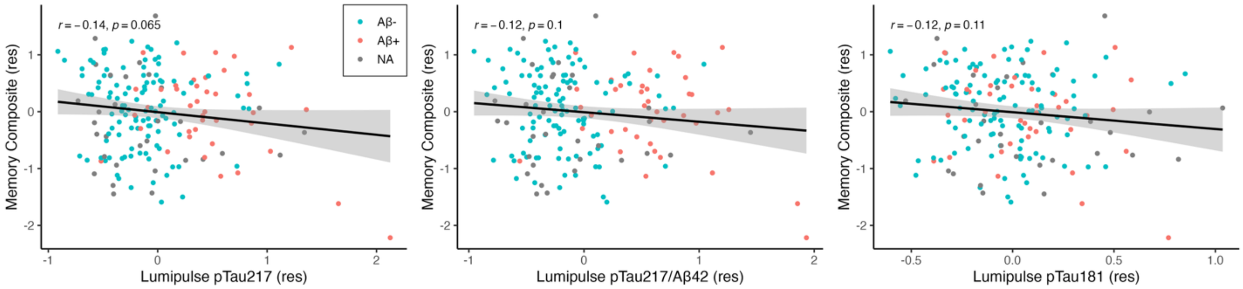


**Supplementary Figure 4.** Association between Lumipulse plasma biomarkers and memory. Residuals controlling for age, sex, and education are plotted. Points are colored by Aβ status defined by CSF Aβ42/Aβ40 or Aβ PET. Scatterplots show regression lines, 95% confidence intervals (shaded area), and Pearson's correlation (*r*).

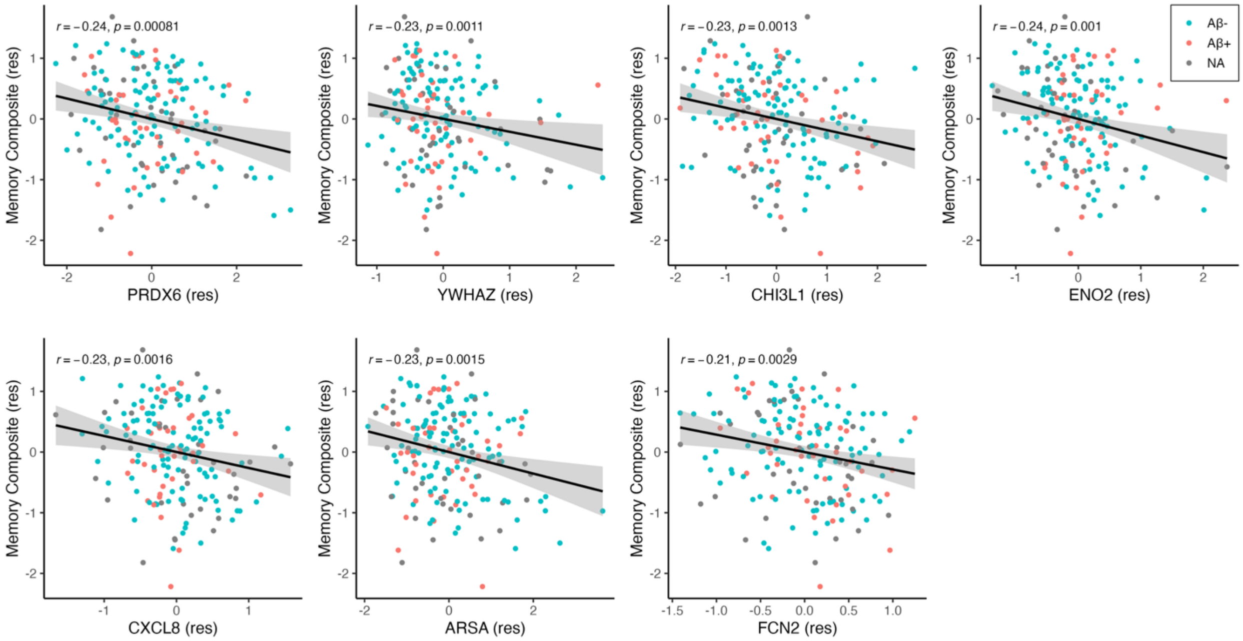


**Supplementary Figure 5.** Association between NULISAseq targets and memory. Residuals controlling for age, sex, and education are plotted. Points are colored by Aβ status defined by CSF Aβ42/Aβ40 or Aβ PET. Scatterplots show regression lines, 95% confidence intervals (shaded area), and Pearson's correlation (*r*).
